# They stumble that run fast: the economic and COVID-19 transmission impacts of reopening industries in the US

**DOI:** 10.1101/2020.06.11.20128918

**Authors:** Marita Zimmermann, Amy E Benefield, Benjamin M Althouse

**Affiliations:** Institute for Disease Modeling, Bellevue, WA; University of Washington, Seattle, WA; University of Colorado, Boulder, CO; New Mexico State University, Las Cruces, NM

## Abstract

COVID-19 has laid bare the United States economically and epidemiologically. Decisions must be made as how and when to reopen industries. Here we quantify economic and health risk tradeoffs of reopening by industry for each state in the US. To estimate total economic impact, we summed income loss due to unemployment and profit loss. We assess transmission risk by: (1) workplace size, (2) human interactions, (3) inability to work from home, and (4) industry size. We found that the industry with the highest estimated economic impact from COVID-19 was manufacturing in 40 states; the industry with the largest transmission risk index was accommodation and food services in 41 states, and the industry with the highest economic impact per unit of transmission risk, interpreted as the value of reopening, was manufacturing in 37 states. Researchers and decision makers must work together to consider both health and economics when making tough decisions.

> *O, let us hence; I stand on sudden haste*.
>
> *Wisely and slow; they stumble that run fast*.
>
> -William Shakespeare, [Romeo and Juliet](https://en.wikiquote.org/wiki/Romeo_and_Juliet), Act II, scene 3

## Background

The first confirmed case of COVID-19 in the United States was a travel-associated case in Snohomish County, Washington screened on January 19^th^, 2020 (1). As of June 6^th^, the United States has recorded 1,917,080 cases and 109,702 deaths (2). On February 29^th^, Governor Jay Inslee declared a state of emergency in Washington, soon after which companies began urging employees to work from home if possible (3). On March 13^th^, President Trump declared a national emergency (4). On March 23^rd^, eight states had statewide stay-at-home orders in place, and by April 7^th^ that had increased to 42 states (5). Stay-at-home-orders began to expire as early as April for a few states, with most lasting in to May and a few in to June (5). All 50 states have begun reopening, though approaches to doing so vary(5).

While shutdown measures have been necessary to control the spread of COVID-19, they also take a toll on the economy: unemployment in the U.S. is over 13%, with the highest rate in Nevada, at more than 24% (6). However, heedlessly reopening the economy could lead not only to drastic increases in COVID-19 cases, but also to even greater economic downturn due to added health burden and consumer risk (7). Part of planning to reopen the economy will be carefully considering which businesses to reopen and when to do so.

The economic impacts of the shutdown have not been uniform across industries; therefore, reopening will have differential impacts for different industries. Unemployment has centered on industries that rely on in-person interactions, specifically those that cannot be done with remote work such as restaurants and in-person retail. Ideally, leaders will target reopening industries that have experienced large unemployment or overall economic distress due to COVID-19, however, it is also important to consider the transmission risks in each industry. Workplaces that require significant close contact with large numbers of people or utilize work that cannot be done remotely have a higher likelihood of increased transmission than others. Here, we define reopening of an industry to indicate that should stay-at-home orders be lifted, employees able to work from home would continue to do so, and safety interventions such as hand-washing and mask wearing would continue. In this analysis we aim to quantify the tradeoffs between economic impact and health risks for economic reopening by industry for each state in the US.

## Methods

We aimed to evaluate (1) the economic impact of the shutdown and (2) the relative transmission risk of reopening industries in each state. Industries were categorized by the North American Industry Classification System (NAICS) (8). There are many sub-industries and business types within the NAICS industry groupings with economic impacts and transmission risks that may not be accurately reflected in industry-level data. The fact that this report reflects only industry-level data is a limitation. For all data sources referred to below, see Appendix 1 for details. We chose to compare Washington and California in the examples below, due to the first reported case occurring in Washington state and the relevant comparison of California due to proximity and large economy.

### Economic Impact

To estimate total economic impact by industry (*Econ*), we summed income loss due to unemployment (*Unemp*) and profit loss (*Prof*) as follows:

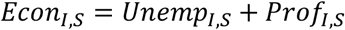

where *Econ* is the total economic impact in industry *I* in state *S*. All costs were converted to 2019 USD.

To estimate income loss due to unemployment by industry and state, *Unemp*_*I,S*_, we used continued unemployment claims from April 2020 (*U*_*I,S*_) to calculate the industry distribution of unemployment due to COVID-19. Continued unemployment claims are reported for each state by industry once a month, representing continued claims the week of the 19^th^, or people who were unemployed the week of the 12^th^of each month (9). We then applied the industry unemployment proportions to the total unemployment due to COVID-19 in each state. Total unemployment due to COVID-19 in each state was calculated as the difference between weekly unemployment percent, *σ*_*S*_, the week of May 23^rd^ and the average weekly unemployment percent for all of 2019 (6). This rate was applied to the total civilian employed population age 16 and older in the state, *N*_*S*_ (10), and finally average weekly wage in that industry *ω*_*S,I*_ (11).

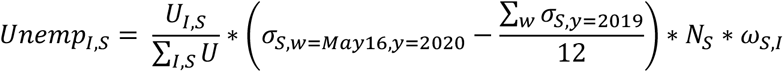

To estimate profit loss, *Prof*_*I,S*_, we first found the percent decrease in sales likely attributed to Covid-19. Using SafeGraph weekly mobility data (12), we calculated the percent decrease in customer interactions, *μ*, in each industry from the week of March 30^th^, 2020 (the lowest point of mobility) to the equivalent week in 2019. SafeGraph is a data company that aggregates anonymized location data from numerous applications in order to provide insights about physical places. To enhance privacy, SafeGraph excludes census block group information if fewer than five devices visited an establishment in a month (two devices in a week) from a given census block group. Customer interactions include any interaction lasting less than four hours (12). We assumed that any job that can be done from home (*δ*_*I*_) would not be subject to profit loss (13). For jobs that cannot be done from home (1 − *δ*_*I*_), we calculated profit as the weekly revenue, *ρ*_*I,S*_ (14), multiplied by the expected profit margin, *θ*_*I*_. (15).

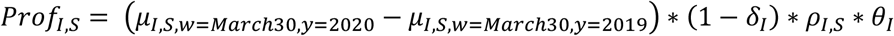

### Transmission Risk Index

We calculated a transmission risk index (*T*_*S,I*_) based on a summation of four factors: (1) workplace size: average number of employees per individual workplace (*P*_*S,I*_), normalized 0-1 with 0 representing the smallest number of employees per workplace and one representing the largest; (2) human interactions: expected number of human interactions that would resume if the industry were reopened (*H*_*S,I*_), normalized 0-1; (3) inability to work from home: percent of jobs that cannot be done from home in each industry (*δ*_*I*_), normalized 0-1; and (4) industry size: total number of employees in each industry in each state(*N*_*S,I*_), normalized 0-1. The transmission risk index is calculated as follows:

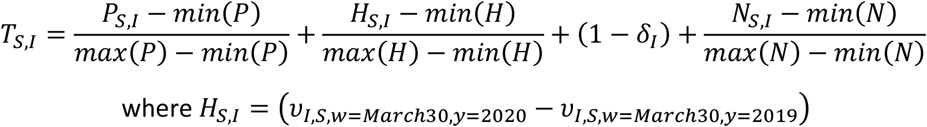

and where *υ*_*I,S,w,y*_ is the number of human interactions in industry *I*, state *S*, during week *w* of year *y*.

This risk index is intended to quantify potential transmission risk differences between industries. It cannot be used to directly predict cases or disease burden.

We hypothesized that a workplace building with more employees would be more likely to facilitate higher levels of transmission. For this factor, we found the mean number of employees per establishment in each industry in each state and normalized the result from zero to one (16). This data source does not include the physical size of each workplace (e.g., warehouse workers may have less frequent physical contact than those in office buildings), which is a limitation of this approach.

In addition to potential interactions between employees, we aimed to quantify a factor for total human interactions in each industry that have been eliminated due to social distancing and shutdown policies. These interactions would be expected to restart if the industry were reopened. We used SafeGraph weekly mobility data to calculate the difference in total interactions *υ*_*I,S,w,y*_ between the week of March 30^th^, 2020, which was the lowest point of mobility, to the equivalent week in 2019 for each industry, and we assigned interaction scores on a scale from zero to one, with zero representing the fewest interactions and one representing the most (12).

Additionally, we included a factor for the percent of jobs that cannot be done from home for each industry and included a factor for the size of the industry, measured by total number of employees in each state, normalized from zero to one (10,13). The transmission risk index assumes that if the industry was reopened, those who can do their job from home would continue to do so.

## Results

In California, for example, the top three industries for economic impact due to COVID-19 shutdowns were (1) manufacturing, (2) healthcare and social assistance, and (3) retail trade, compared to (1) manufacturing, (2) construction, and (3) healthcare and social assistance in Washington (Figure 1). For results for all states see Appendix 2 and https://idmodresearch.shinyapps.io/industry_state/.

**Figure 1.**
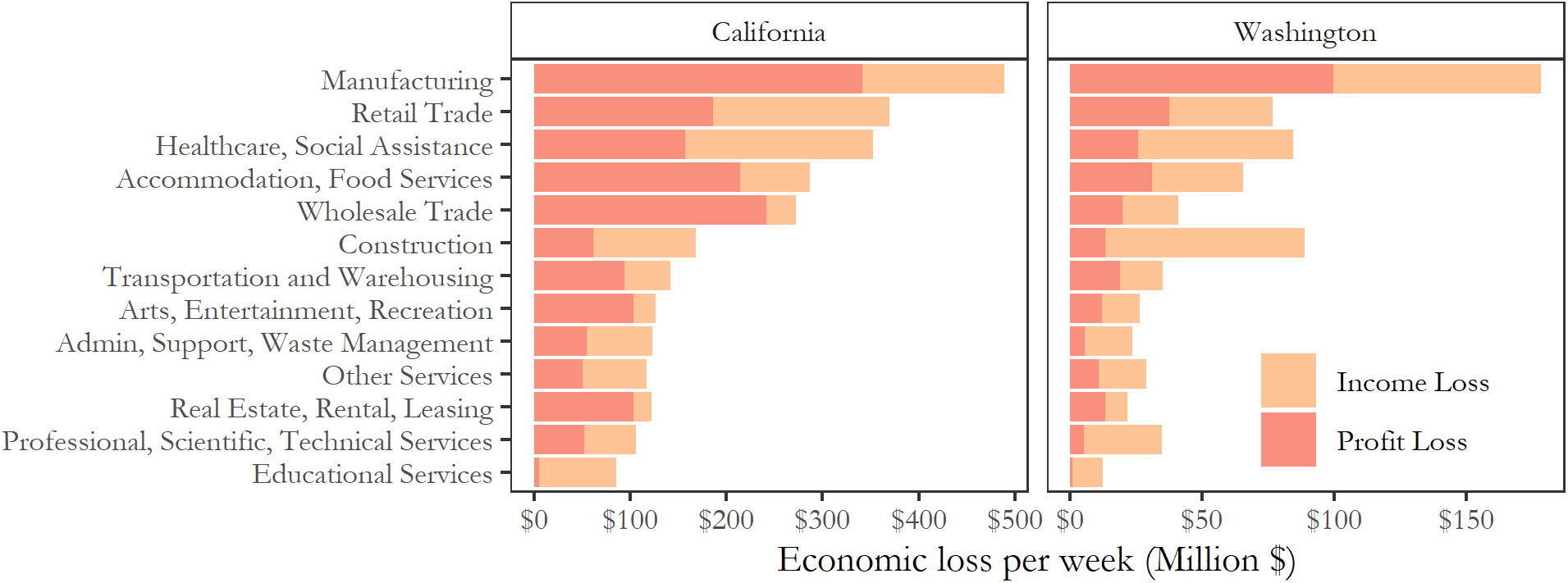
Economic impact from income loss due to unemployment and profit loss attributed to COVID-19 in California and Washington. The highest estimated economic impact due to COVID-19 in was in manufacturing. Note that X-axis scales differ between figure panels.

After summing the four factors of workplace size, human interactions, inability to work from home, and industry size, in both California and Washington, the industries with the highest transmission risk index were (1) accommodation and food services, (2) retail trade, and (3) healthcare and social assistance (Figure 2). For results for all states see Appendix 2 and https://idmodresearch.shinyapps.io/industry_state/.

**Figure 2.**
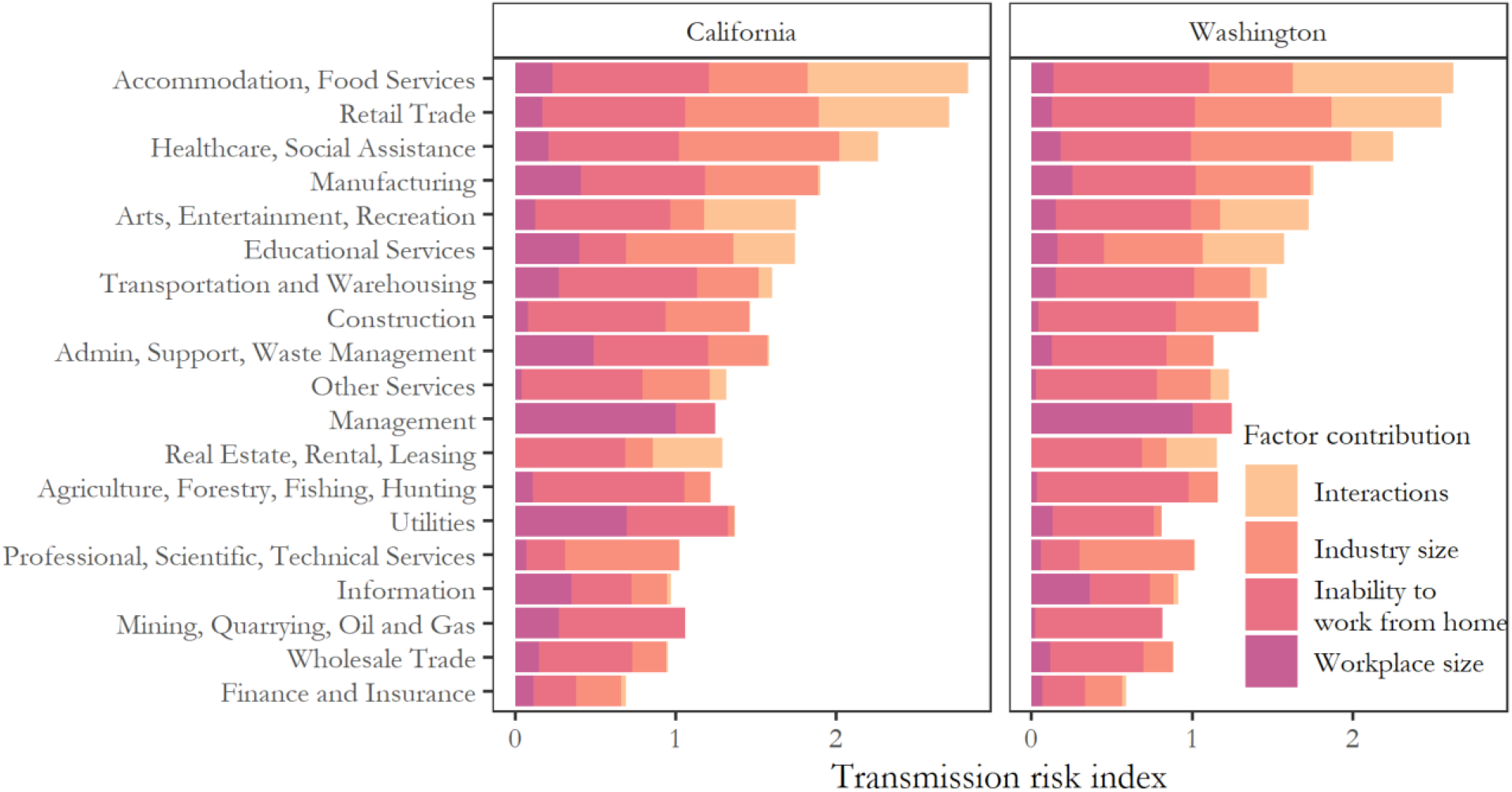
Total transmission risk index calculated as a sum of contributions from four factors in California and Washington. The four factors are: (1) workplace size: normalized number of employees per individual workplace in each industry, (2) interactions: normalized number of human interactions that would re-start if industry was reopened, (3) inability to work from home: percent of jobs that cannot be done from home in each industry, and (4) industry size: normalized number of employees in each industry in Washington. The largest transmission risk index was found in accommodation and food services.

In examining economic impact and transmission risk simultaneously for reopening purposes, we should target industries for reopening that have a high economic impact and low transmission risk. In terms of tradeoffs of economic losses from the shutdown versus the health risk of reopening, industries toward the top left of the Figure 3 would have the most economic gain for minimum health risk. The industry with the highest economic impact per unit of transmission risk was wholesale trade in California and manufacturing in Washington (Figure 3).

**Figure 3.**
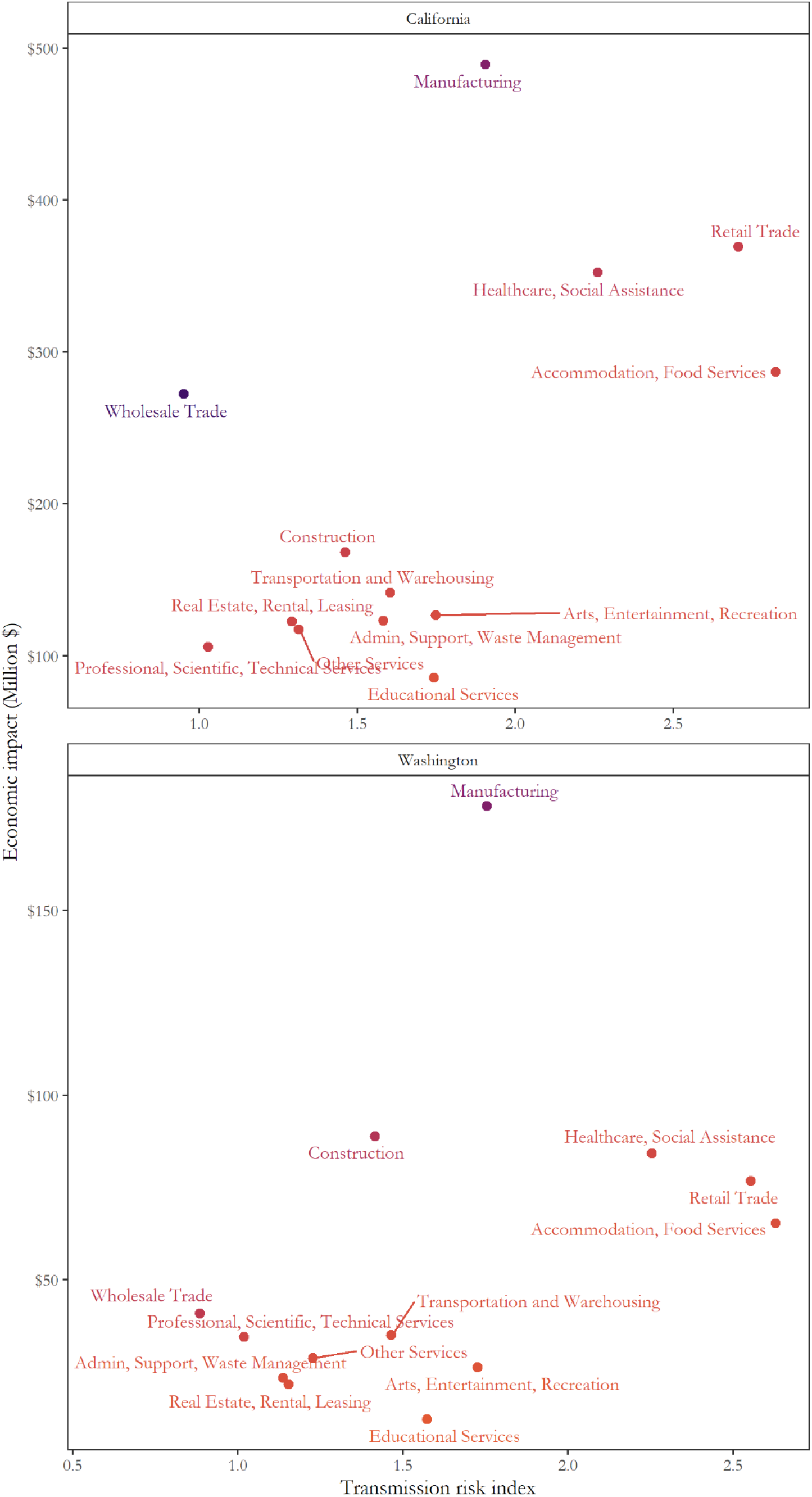
Estimated weekly economic impact of COVID-19 and transmission risk index for each industry in California and Washington. Wholesale trade and manufacturing had the highest economic impact relative to their transmission risk in California and Washington, respectively. Therefore, they should be targeted for earlier reopening. Colors represent the gradient of highest economic gain per unit of transmission risk (purple) to lowest (orange). Note that axis scales differ between figure panels.

We found that the industry with the highest estimated economic impact due to COVID-19 was manufacturing in 40 states; accommodation and food services in six states (AZ, CO, FL, HI, NV, and NY); healthcare and social assistance in three states (AK, MD, and RI); and wholesale trade and other services (which includes repair and maintenance; personal and laundry services; religious, grantmaking, civic, professional, and similar organizations; and private households) in one state each (NJ and DC, respectively) (Figure 4, top left). We found that the industry with the largest transmission risk index was accommodation and food services in 41 states; retail trade in five states (CT, ME, NH, NJ, and UT); healthcare and social assistance in three states (ND, SD, and VT); and manufacturing and educational services each in one state (IN and IA, respectively) (Figure 4, top right). Finally, we found that the industry with the highest economic impact per uint of transmission risk was manufacturing in 37 states; wholesale trade in ten states (AZ, CA, CO, CT, FL, MA, MD, NJ, NY, and RI); accommodation and food services in two states (HI and NV); and construction and other services each in one state (AK and DC, respectively) (Figure 4, bottom left). These can be interpreted to have the highest value of reopening.

**Figure 4.**
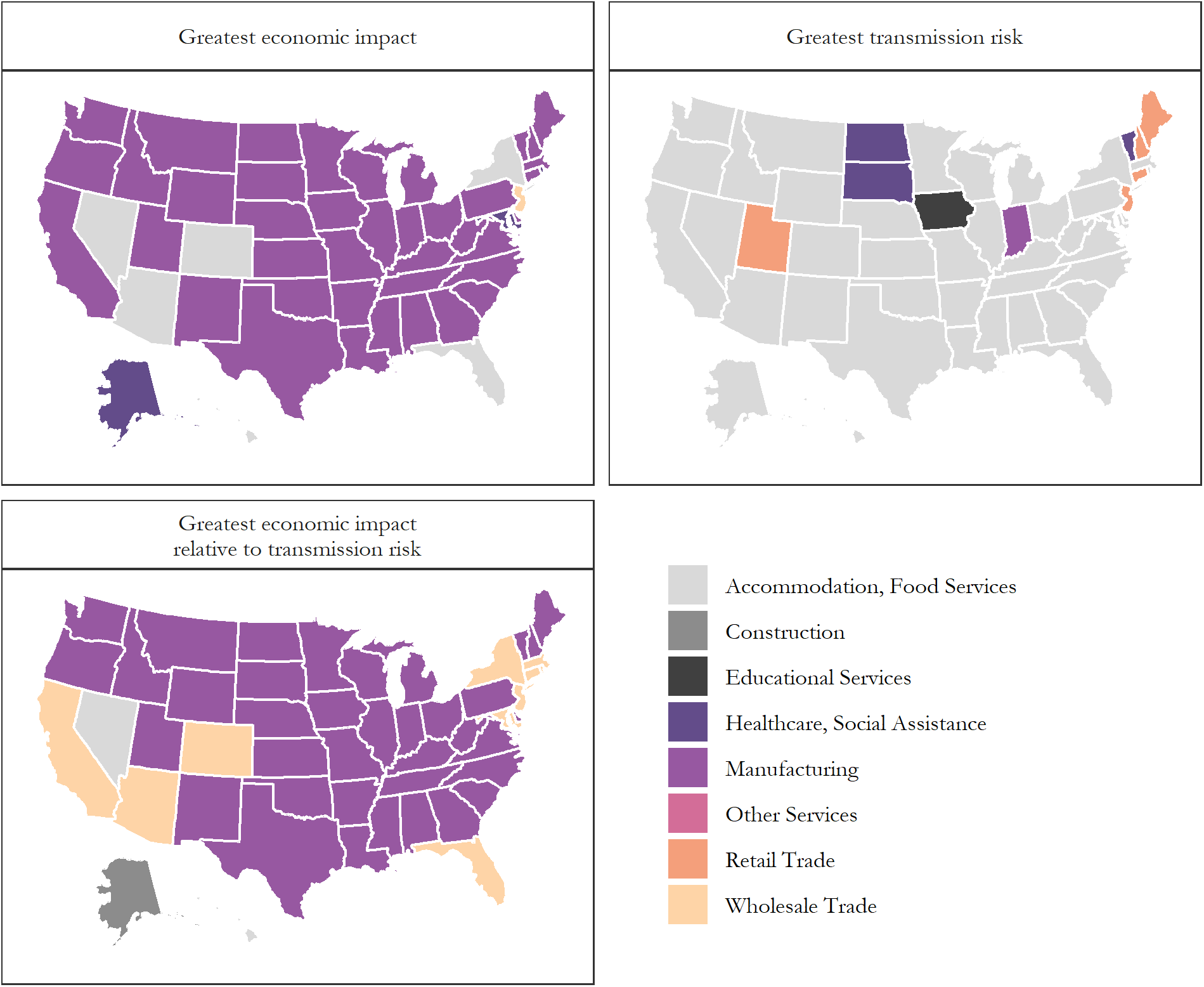
For each state, industry with the highest economic impact of COVID-19 shutdowns (top left), highest transmission risk index (top right), and highest economic impact per transmission risk index, or value of reopening (bottom left)

## Discussion

While reopening states and returning individual livelihoods to the millions of unemployed Americans is of key importance, it must be done in balance to limit further spread of COVID-19, which has claimed over 100,000 American lives, sickened nearly 2 million, and ravaged healthcare systems across the US. In estimating the economic impact due to COVID-19 alongside the relative transmission risk in each industry by state, we find that the manufacturing industry would be a good target for early reopening in the majority of states. This is driven by the high economic impact of reopening this industry with relatively lower COVID-19 transmission risk. However, it is important to note that high reopening value does not necessarily imply low transmission risk. Indeed, in Nevada, Hawaii, and Indiana, the industry with the highest value for reopening also has the highest transmission risk index. For this reason, each state must weigh priorities carefully in establishing reopening strategies.

Use of transmission mitigation strategies will be essential to ensure a safe reopening of the economy and determine industry reopening readiness. Due to the nature of the industry, accommodation and food services, for example, presents the highest transmission risk in most states. This risk, however, may be mitigated though strategies such as capping seating capacity and robust personal hygiene, in which case restaurants would be a key target for early reopening. Furthermore, the mitigation tactics – and the ease to which they are implemented and enforced – will need to be thoughtfully considered, as they will vary significantly between- and within-industries. For instance, within the arts, entertainment, and recreation industry, social distancing may be appropriate for both movie theaters and indoor concert venues but it might only be enforceable in the movie theater setting. This may indicate that movie theaters are a target for early reopening but that concert venues remain closed for the time being. Similarly, transmission risk is likely lower in large manufacturing facilities where employees have little interaction with each other or those that take place outdoors compared to smaller or indoor operations.

The healthcare industry has high economic impact and high transmission risk in several states and should be given careful consideration for several reasons. First, the industry is essential, so it has not been fully closed. Instead, we consider reopening of the health care industry to mean restarting non-emergent and elective care. We must consider transmission risk more carefully in this population: On one hand, healthcare settings may have increased transmission risk due to likelihood of interactions with infected people; on the other, they are best equipped to prevent the spread of infections (both in training and in personal protective equipment) and have frequent testing and symptom screening of healthcare employees.

Similarly, we must give careful thought to the education industry. First, for most settings within education, economic gain may not be the primary goal. This analysis does not incorporate the future effects of limited or lower quality education due to shelter-in-place policies. Additionally, our analysis is based on data assuming that teachers can work from home, which may not be the case for effective education, particularly for young children. We also do not consider the disproportionate effects of closing schools on marginalized and vulnerable populations independently, or the effects on other industries of parents having limited childcare options during work hours, which is a limitation of this approach (17–19). For these reasons, we recommend that education not be directly compared to other industries, and instead be given special consideration for reopening purposes. Routine COVID-19 testing (weekly, or even semiweekly) and frequent symptom screening may help schools reopen safely; but cheaper PCR tests or pooling of specimens may have to be implemented in the face of limited resources in schools.

This work is meant to inform initial discussions regarding the reopening of industries. While it illuminates high-level industry economic impacts and transmission risks, there are a host of other considerations vital to the policymaking processes. Questions remain regarding the social, cultural, and mental health impacts of industry closures that may outweigh risks of reopening. Data is newly surfacing on how the shutdown has disproportionately affected communities of varying socio-economic status. We need to understand this better and ensure that burden relief reaches the most affected. These, among many other, questions and considerations need to be a part of the process with which policymakers develop policy and redefine the road back to an open economy. Our work emphasizes that the reopening of states should not be done in haste, nor should it be motivated by political agenda. We can and should use data-driven approaches to control this unprecedented public health risk. Lives are at stake, both due to COVID-19 disease itself and dire economic consequences for millions of Americans. Researchers and decision makers must work together to consider both health and economics when making tough decisions.

## Data Availability

All data used in this analysis is publicly available.

## References

1. Novel Coronavirus Outbreak 2020 (COVID-19)?:: Washington State Department of Health [Internet]. [cited 2020 May 18]. Available from: https://www.doh.wa.gov/emergencies/coronavirus

2. Johns Hopkins University (JHU). COVID-19 Dashboard by the Center for Systems Science and Engineering (CSSE) [Internet]. [cited 2020 Jun 6]. Available from: https://gisanddata.maps.arcgis.com/apps/opsdashboard/index.html#/bda7594740fd40299423467b48e9ecf6

3. COVID-19 Resources and Information | Governor Jay Inslee [Internet]. [cited 2020 May 14]. Available from: https://www.governor.wa.gov/issues/issues/covid-19-resources

4. Proclamation on Declaring a National Emergency Concerning the Novel Coronavirus Disease (COVID-19) Outbreak | The White House [Internet]. [cited 2020 Jun 6]. Available from: https://www.whitehouse.gov/presidential-actions/proclamation-declaring-national-emergency-concerning-novel-coronavirus-disease-covid-19-outbreak/

5. See How All 50 States Are Reopening - The New York Times [Internet]. [cited 2020 Jun 6]. Available from: https://www.nytimes.com/interactive/2020/us/states-reopen-map-coronavirus.html

6. United States Department of Labor Employment & Training Administration. Unemployment Insurance Weekly Claims Data [Internet]. 2020 [cited 2020 Jun 6]. Available from: https://oui.doleta.gov/unemploy/claims.asp

7. Reopening U.S. economy amid coronavirus too soon could cause “double-dip” recession | Fortune [Internet]. [cited 2020 May 18]. Available from: <https://fortune.com/2020/05/11/us-economy-reopen/>

8. United States Census Bureau. North American Industry Classification System (NAICS) Main Page.

9. Department of Labor. Unemployment claims table AR203 [Internet]. [cited 2020 Jun 6]. Available from: https://oui.doleta.gov/unemploy/chariu.asp

10. Table B24050: Industry by Occupation for the Civilian Population - Census Reporter [Internet]. [cited 2020 Jun 6]. Available from: https://censusreporter.org/tables/B24050/

11. Table B24031: Industry by Median Earnings for the Civilian Population - Census Reporter [Internet]. [cited 2020 Jun 6]. Available from: https://censusreporter.org/tables/B24031/

12. SafeGraph. SafeGraph COVID-19 Data Consortium [Internet]. [cited 2020 Jun 6]. Available from: https://www.safegraph.com/covid-19-data-consortium

13. Dingel JI, Neiman B. How Many Jobs Can be Done at Home? [Internet]. 2020 [cited 2020 May 14]. Available from: https://bfi.uchicago.edu/wp-content/uploads/BFI_White-Paper_Dingel_Neiman_3.2020.pdf

14. Economic Census [Internet]. [cited 2020 Jun 6]. Available from: https://www.census.gov/programs-surveys/economic-census.html

15. Damodaran A. Operating and Net Margins [Internet]. [cited 2020 May 14]. Available from: http://pages.stern.nyu.edu/~adamodar/New_Home_Page/datafile/margin.html

16. US Census Bureau. 2017 SUSB Annual Datasets by Establishment Industry.

17. With schools closing across the U.S., kids from low-income families will struggle to learn at home - MarketWatch [Internet]. [cited 2020 May 14]. Available from: https://www.marketwatch.com/story/with-schools-closing-across-the-us-kids-from-low-income-families-will-struggle-to-learn-at-home-2020-03-15

18. Coronavirus crisis: LA County sees spike in reported domestic violence reports amid pandemic - ABC7 Los Angeles [Internet]. [cited 2020 May 14]. Available from: <https://abc7.com/domestic-violence-coronavirus-stay-at-home-pandemic/6119703/>

19. Coronavirus: Rural California children face challenges after school closures - Los Angeles Times [Internet]. [cited 2020 May 14]. Available from: https://www.latimes.com/california/story/2020-04-22/coronavirus-rural-schools-california

